# Cohort Profile: The Birhan Health and Demographic Surveillance System

**DOI:** 10.1101/2021.01.21.21249989

**Authors:** Delayehu Bekele, Bezawit Mesfin Hunegnaw, Chalachew Bekele, Kimiko Van Wickle, Fisseha Tadesse, Frederick G. B. Goddard, Yahya Mohammed, Sarah Unninayar, Grace J. Chan

**Affiliations:** Department of Obstetrics and Gynecology, St. Paul’s Hospital Millennium Medical College, Addis Ababa, Ethiopia; Department of Pediatrics and Child Health, St. Paul’s Hospital Millennium Medical College, Addis Ababa, Ethiopia; Birhan HDSS, St. Paul’s Hospital Millennium Medical College, Addis Ababa Ethiopia; Department of Epidemiology, Harvard T.H. Chan School of Public Health, Boston, MA, USA; Department of Obstetrics and Gynecology, Debire Birhan Referral Hospital, Debire Birhan, Ethiopia; Division of Medicine Critical Care, Boston Children’s Hospital, Department of Pediatrics, Harvard Medical School, Boston, MA, USA

**Keywords:** Birhan HDSS, health and demographic surveillance, pregnancy and birth, mortality, morbidity, maternal and child health

## Abstract

The Birhan health and demographic surveillance system (HDSS) was established to determine the magnitude and causes of morbidity and mortality in Ethiopia among pregnant and postpartum women and children. Located in the North Shewa Zone of the Amhara Region, the site includes 16 kebeles (villages) and spans two woredas (districts), Angolela Tera and Kewet/Shewa Robit. The initial census was conducted in May 2018, and core demographic and health events are updated every three months. The site has 18,933 households and a population of 77,766. Over 82.6% of the population is rural.

During the baseline census, all households were geocoded; household members were enumerated; and data were collected on sociodemographic status, housing construction material, economics and asset ownership, care seeking behaviors, and water and sanitation access. During each subsequent round, data on key demographic events such as births, deaths, marital status changes, in-migration, or out-migration were collected for all enumerated household members. Pregnancy surveillance was conducted among married women of reproductive age 15-49 using a pregnancy screening questionnaire and confirmed by urine pregnancy tests. Morbidity and immunization status data for children aged under two years were collected during each round.

## Why was the HDSS set up?

Although there has been significant progress in reducing maternal and child mortality over the past few decades, the rates remain high globally. Nearly 94% of maternal mortality occurs in low-middle income country (LMIC) settings and arises from largely preventable causes(1). In Ethiopia, the maternal mortality rate is 412/100,000 live births, amounting to 14,000 maternal deaths each year(2, 3). Sub-Saharan Africa is the region with the highest under-five mortality and Ethiopia is one of five countries which contributed to more than half of the global under-five deaths in 2018(4). Ethiopia is one of 10 countries accounting for more than half of global neonatal deaths with 80,000 deaths each year(5). Complete, timely, and accurate data are essential to understand the causes and underlying risk factors of maternal and child morbidity and mortality. These data and the resulting knowledge generated are subsequently key to the development of effective interventions to improve health outcomes. Currently, health data in Ethiopia remain limited in scope and quality, and data collected at community and health facility levels are not linked.

The Birhan HDSS (Birhan meaning ‘light’ in Amharic), located near Debre Birhan, Amhara Region, was established as a sentinel field site to generate evidence on maternal and child health in Ethiopia. The Birhan HDSS collects socioeconomic, demographic, and health data in a defined catchment population to improve morbidity and mortality surveillance. Nested within the Birhan HDSS is a pregnancy and birth cohort. Together, the Birhan HDSS and cohort provide detailed data on time trends in maternal and child health to better understand the etiologies of disease and predict risk. The platform was designed to support nested studies to assess the effects of exposures and interventions on outcomes. Information on vital events such as births and deaths build the foundation for equitable health policies.

## Where is the HDSS area?

The Birhan HDSS was established in Ethiopia, a landlocked country in East Africa bordering Djibouti, Eritrea, Kenya, Somalia, South Sudan, and Sudan. The HDSS is located in North Shewa Zone, one of the ten zones in Amhara Region (Figure 1). The zone is bordered on the south and west by Oromia Region, on the north and northeast by South Wollo Zone and Oromia Zone of Amhara Region, and on the east by Afar Region(6). The capital city of the zone, Debre Birhan, is located 130 km north of Addis Ababa, the country’s capital.

**Figure 1.**
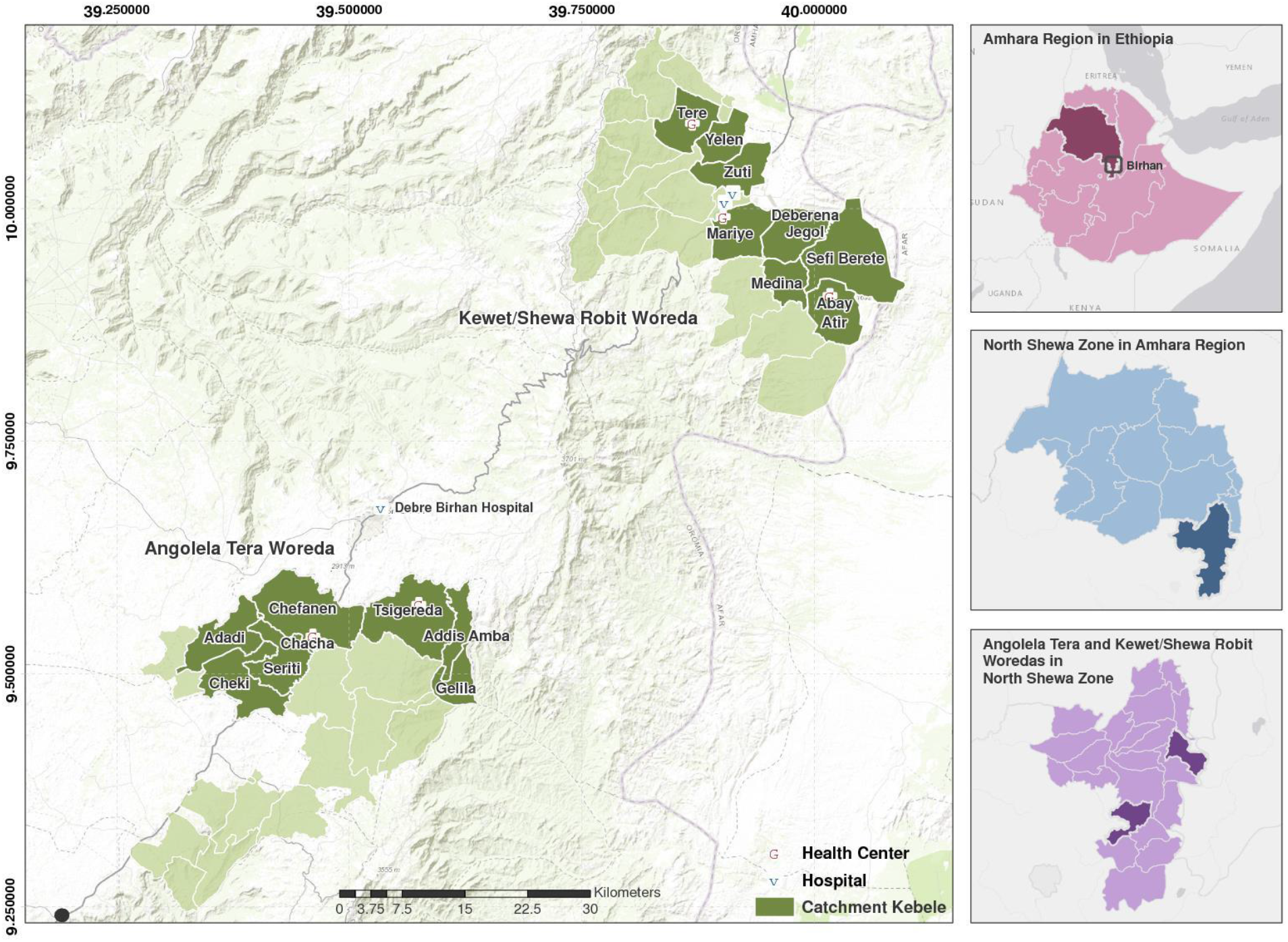
Map of the Birhan HDSS

To select the site, the study team conducted rapid assessments of surrounding areas within 300 km of Addis Ababa to evaluate based on a set of selection criteria including the distribution of diseases, representative mix of rural and urban kebeles, highland and lowland topography, diversity of ethnicities, and safety. We also considered recommendations from the zonal health bureau.

The study site consists of two districts—Angolela Tera and Kewet/Shewa Robit - covering a total area of 636 sq km. Each district has eight kebeles (the lowest administration unit). Of the 16 total kebeles, two are urban and the remaining 14 are rural. The study kebeles in Angolela Tera District are located 111 km north of Addis Ababa and are predominantly highland, with an altitude ranging from 1600-3030 m above sea level(7). The monthly minimum average temperature is −0.1°C, maximum average temperature is 22.9°C, and the monthly average rainfall is 25 mm (range 0-120 mm)(8). The Kewet/Shewa Robit study kebeles are located 220 km north of Addis Ababa and are predominantly at lower elevation, with an altitude ranging from 1093-2155 m above sea level(7). The monthly average temperature ranges from a minimum of 10.2°C to a maximum of 36.4°C, and the monthly average rainfall is 23 mm (range 0-80 mm)(8).

Most residents are farmers, with rain-based farming in the Angolela Tera kebeles and irrigation-based and semi-pastoralist type of practices in the Kewet/Shewa Robit kebeles. There are 47 primary and three secondary schools, 53 churches, and 29 mosques in the study area. The HDSS site includes 16 health posts, five health centers, one primary hospital, and one private hospital. Debre Birhan Hospital serves as a referral site for all facilities in the study catchment.

## What does it cover now?

Following an open prospective cohort design, the Birhan HDSS routinely collects information on health and demographic events including migration (i.e., in-migration, out-migration, and internal moves), births and deaths, marital status, household wealth, types of employment, water supply and sanitation access, and child health. The Birhan HDSS conducts pregnancy surveillance among women 15-49 years of age. Pregnant women and children under two are enrolled into a pregnancy and birth cohort with more frequent follow-ups (both at home and at health facilities) to assess exposures, outcomes, disease symptoms, and anthropometrics.

## Who is covered by the HDSS and how often have they been followed up?

The Birhan HDSS covers all eligible residents in the defined catchment area. To identify and enumerate eligible residents, a baseline household census was conducted from May to August 2018. The initial census identified all houses - defined as a structure made from permanent building material. Houses were enumerated using a unique 10-character identification system. After obtaining consent, a numbering plate was affixed to each door. A household was defined using the definition as members who “share a common pot” for cooking(9). Members of the household were defined as individuals having a dwelling within the household and having lived in the household for at least three months prior to the survey. This included students with temporary residence elsewhere but returning home for weekends or for vacation.

Each household and household member has a unique identification number to allow for longitudinal follow-up. New individuals and households are added to the study through birth or in-migration into the study area. The population is young with a broad-based population pyramid (Figure 2). The study population is visited every three months at home and the health and demographic data are updated by trained study data collectors.

**Figure 2:**
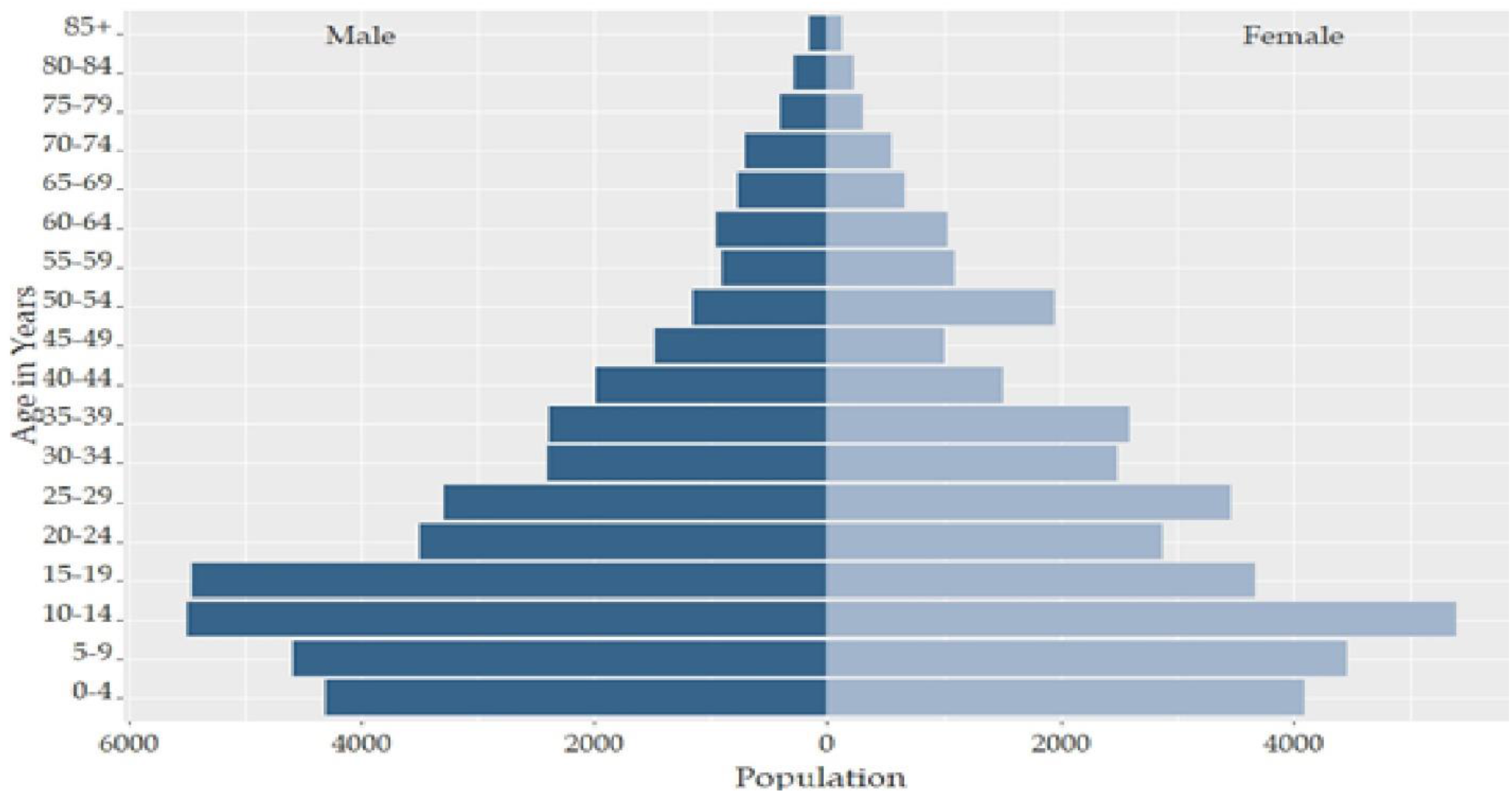
Population pyramid of Birhan HDSS by age and sex, January - December 2019

## What has been measured and how have the HDSS databases been constructed?

During the baseline census, data collectors went door-to-door in each kebele to collect data on household location (including GPS coordinates), sociodemographic status, housing construction material, economics and asset ownership, care seeking behaviors, water and sanitation access and practices, prior pregnancies, and birth histories for the past year (Table 1). During each subsequent round of house-to-house data collection every three months, data on key demographic events such as births, deaths, marital status changes, in-migration, and out-migration are collected for all enumerated household members. Weight, height, and mid upper arm circumference (MUAC) measures are collected for women of reproductive age (15-49). Data collectors conduct pregnancy surveillance among married women of childbearing age by asking pregnancy screening questions. Urine pregnancy tests are done for women who screened positive (10). To assess the prevalence of child morbidities during routine surveillance, data collectors ask mothers/caretakers to recall clinical symptoms of diarrhea, pneumonia, and febrile illnesses occurring within the past two weeks(11-15).

**Table 1.**
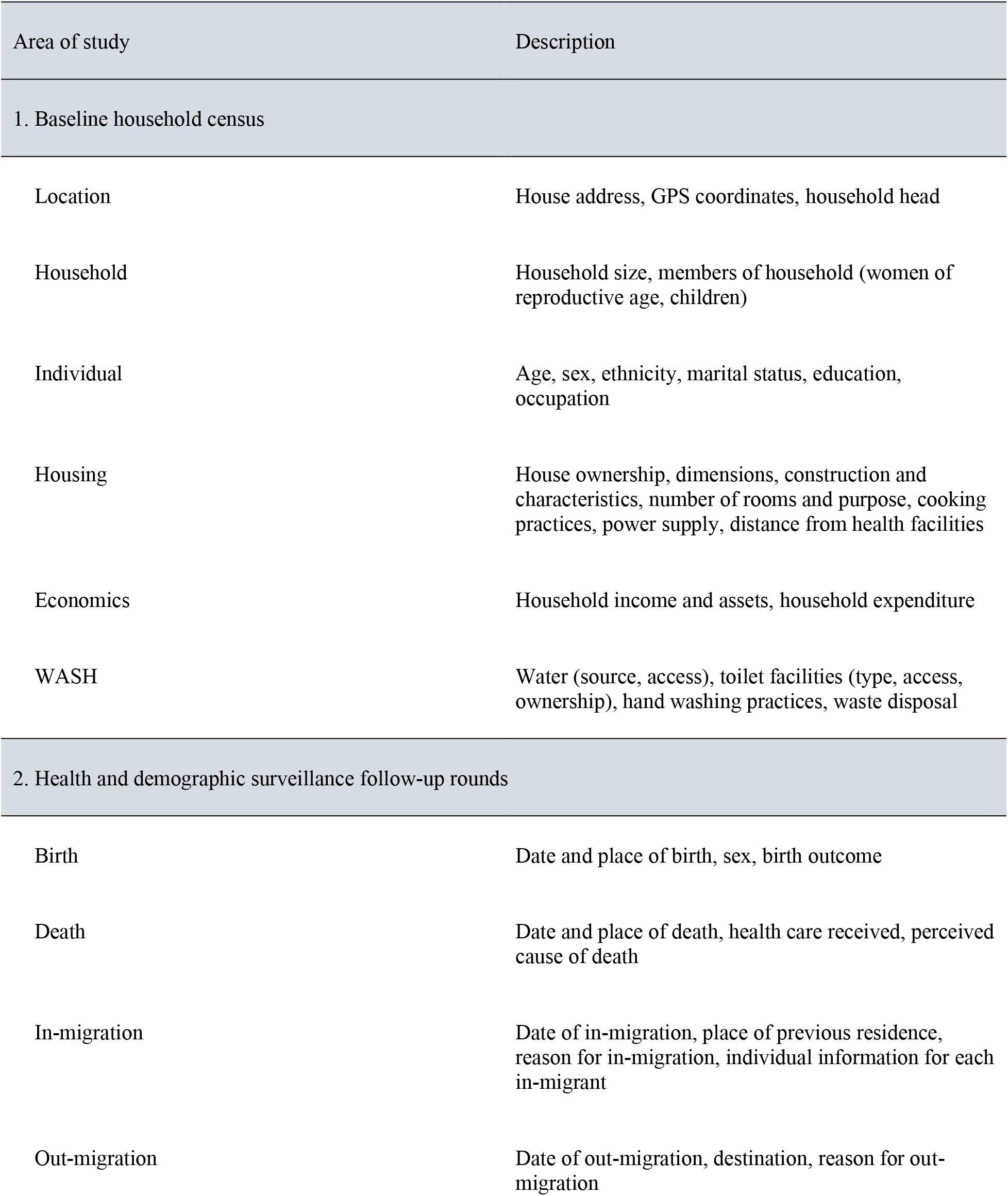
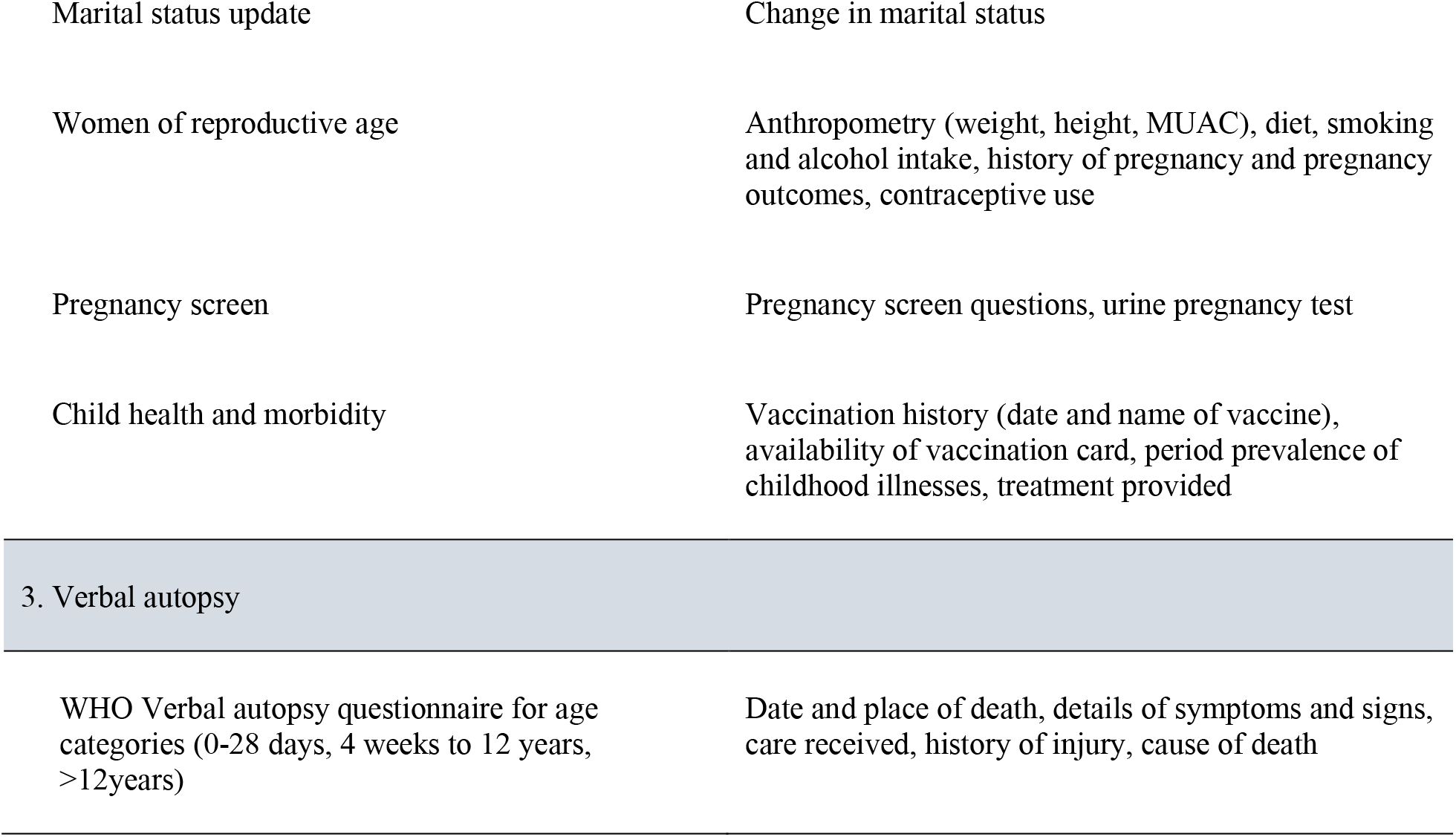
Data collection activities at baseline and follow up rounds of the Birhan HDSS

| Area of study | Description |
| --- | --- |
| 1. Baseline household census |  |
| Location | House address, GPS coordinates, household head |
| Household | Household size, members of household (women of reproductive age, children) |
| Individual | Age, sex, ethnicity, marital status, education, occupation |
| Housing | House ownership, dimensions, construction and characteristics, number of rooms and purpose, cooking practices, power supply, distance from health facilities |
| Economics | Household income and assets, household expenditure |
| WASH | Water (source, access), toilet facilities (type, access, ownership), hand washing practices, waste disposal |
| 2. Health and demographic surveillance follow-up rounds |  |
| Birth | Date and place of birth, sex, birth outcome |
| Death | Date and place of death, health care received, perceived cause of death |
| In-migration | Date of in-migration, place of previous residence, reason for in-migration, individual information for each in-migrant |
| Out-migration | Date of out-migration, destination, reason for out-migration |
| Marital status update | Change in marital status |
| Women of reproductive age | Anthropometry (weight, height, MUAC), diet, smoking and alcohol intake, history of pregnancy and pregnancy outcomes, contraceptive use |
| Pregnancy screen | Pregnancy screen questions, urine pregnancy test |
| Child health and morbidity | Vaccination history (date and name of vaccine), availability of vaccination card, period prevalence of childhood illnesses, treatment provided |
| **3. Verbal autopsy** |  |
| WHO Verbal autopsy questionnaire for age categories (0-28 days, 4 weeks to 12 years, >12years) | Date and place of death, details of symptoms and signs, care received, history of injury, cause of death |

To better understand causes of mortality, verbal autopsy (VA) data are collected using tools adopted from standard World Health Organization (WHO) VA questionnaires for stillbirths, neonates, infants, children, and adults(16). A separate dedicated team of verbal autopsy data collectors conduct VA interviews by visiting caregivers of the deceased after a mourning period of six weeks.

The Birhan HDSS uses an electronic data collection system, developed as a customized version of Open Data Kit (ODK), for longitudinal and relational data collection. The system synchronizes data between surveillance data collectors and other nested studies(17). The database is constructed in a three-level hierarchical tier (i.e., individual, household, village). Data quality checks and logic are built into the data entry tools and data system. At the end of each round, error reports are discussed, and data issues resolved before starting the next survey round.

The Birhan HDSS platform also includes a pregnancy and birth cohort to investigate time-varying exposures and associations with maternal and child health outcomes. A dedicated team of maternal and child health data collectors based at study health facilities and in the community enrolls pregnant women identified through pregnancy surveillance into an open pregnancy and birth cohort. During pregnancy and the postpartum period, community and health facility maternal and child health data collectors follow women at home and at facilities based on a schedule detailed in an additional protocol for the cohort study. Pregnancy surveillance includes ultrasound gestational age dating and measurement of adverse pregnancy outcomes, which will be used to develop pregnancy risk stratification models.

## What has it found? Key findings and publications

During the Birhan HDSS baseline census in May 2018, a total of 79,653 individuals and 19,957 households were identified. Quarterly follow up rounds were conducted in January, April, July, and October of 2019. The 2019 midyear population was 77,766 individuals living in 18,933 households (Table 2). It is a predominantly rural population contributing to 82.6% of the total. The average number of individuals per household was 4.1. The median age is 21 years old (IQR 11.0-36.9). Children under 15 years of age account for 36.0% of the population. The male to female ratio is 1.07 with females accounting for 48.2% of the population. This corresponds well with the projected national ratio of 1.09(18).

**Table 2:** Demographic characteristics of Birhan HDSS, 2019

| Characteristics | Number |
| --- | --- |
| Midyear population | 77,766 |
| Number of households | 18,933 |
| Rural population | 82.6% |
| Average household size | 4.1 |
| Sex ratio at birth (male to female) | 0.95 |
| Sex ratio (male to female) | 1.08 |
| Median age (years) | 21.1 |
| Percent aged under 5 years old | 11.0% |
| Percent aged under 15 years old | 36.0% |
| Dependency ratio | 0.72 |
| Young dependency ratio | 0.62 |
| Old dependency ratio | 0.10 |
| Percent women of reproductive age (15-49 years) | 22.0% |

The site has a total fertility rate of 4.8 births per woman (Table 3), which is slightly higher than the national rate of 4.6 births per woman and significantly higher than the Amhara Region-specific rate of 3.7 births per woman(2). This may reflect the rural majority in the study population.

**Table 3:** Vital rates among study population 2019

| Characteristics | Rate |
| --- | --- |
| Early neonatal mortality rate per 1000 live births | 17.2 |
| Neonatal mortality rate per 1000 live births | 27.6 |
| Infant mortality rate per 1000 live births | 42.1 |
| Under-five mortality rate per 1000 live births | 46.8 |
| Crude birth rate per 1000 people | 24.7 |
| Crude death rate per 1000 people | 5.4 |
| Total fertility rate per woman | 5.0 |

The under-five mortality rate of 47 deaths per 1000 live births (Table 3) is lower than the national rate of 55 deaths per 1000 live births reported in the 2019 Ethiopia Mini Demographic and Health Survey (EMDHS) (19). The neonatal mortality rate of 28 deaths per 1000 live births and infant mortality rate of 42 deaths per 1000 live births are comparable to the 30 and 43 deaths per 1000 live births respectively reported in the 2019 EMDHS.

Overall, 39% of the adult population has had some formal schooling. Nearly two thirds of women (65%) had no formal schooling. Of those that have advanced to more than secondary education, less than one third are females.

Most families live in traditional houses made of wood and mud (91%), with dirt floors (81%) and corrugated iron roofing (78%). The majority of households have access to an improved drinking water source (90%), although few have access to piped water in to their plot (18%) and the majority of households access water from public taps (57%)(20). Only 23% of households have access to an improved latrine (Table 4), with most families having access to a pit latrine with a slab (23%) or without a slab (37%)(20).

**Table 4:**
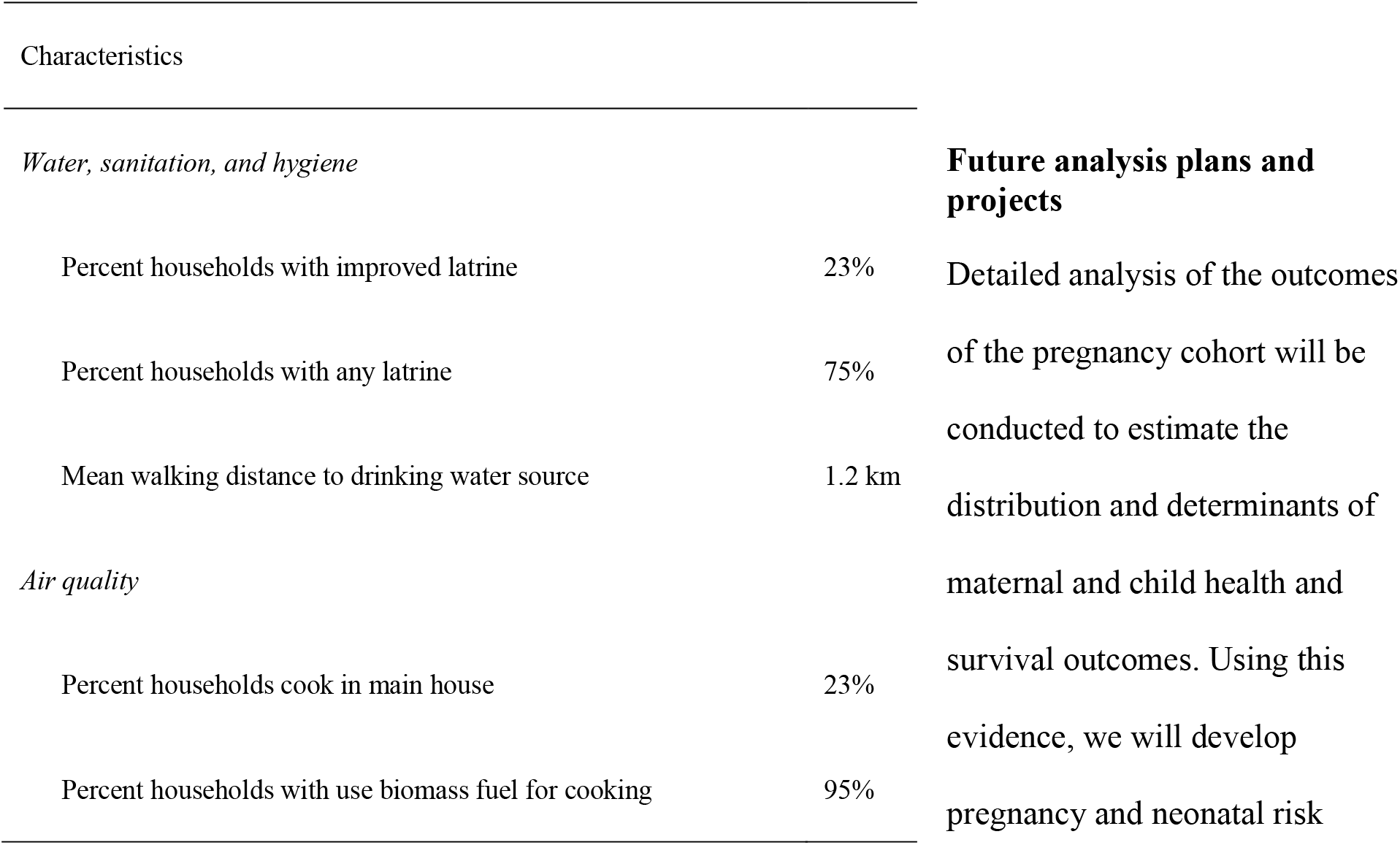
Environmental characteristics

Over two thirds of residents in urban areas have access to household electricity compared to less than a third of rural residents. Two thirds of all households have at least one family member who owns a mobile phone.

### Future analysis plans and projects

Detailed analysis of the outcomes of the pregnancy cohort will be conducted to estimate the distribution and determinants of maternal and child health and survival outcomes. Using this evidence, we will develop pregnancy and neonatal risk stratification algorithms. We hope this will improve clinical diagnosis and care in this low resource setting.

### Strengths and weaknesses

The study area encompasses different ecological conditions with a mix of highland and lowland topography, varying weather patterns, and different farming practices. The environmental, demographic, and economic diversity of the site is representative of the entire Amhara Region. Variables collected are similar to national surveys and other HDSS sites around the world, which will allow for data comparisons.

Quarterly data collection is more frequent than most global HDSS sites. This reduces the lost to follow-up, recall biases, and improves the quality of data. However, frequent rounds of data collection may risk creating community fatigue. To prevent this, we are establishing a community advisory board. This board will collaborate and engage community participation to maintain a good relationship between the study team and the community.

We have developed a de novo online electronic data system that captures longitudinal data from both household and facility visits to accommodate low internet bandwidth. The low internet bandwidth in Ethiopia and the rural study area has created occasional difficulties with data transfer, however the study team has adapted data transfer strategies that are offline and encrypted during internet outages. The study follows an open cohort design to capture the population dynamics in a region of the world that has historically been challenging to conduct such type of studies. As a result, the high in- and out-migration in the area, coupled with the size of the study population, poses analytical challenges to define a steady state population. To calculate rates, a mid-year population was used.

### Data sharing and collaboration

Through the Birhan HDSS platform, data are jointly owned by St. Paul’s Hospital Millennium Medical College (SPHMMC) in Ethiopia and Harvard-Boston Children’s Hospital (BCH) in the United States. Birhan HDSS is open to opportunities for collaborative research both nationally and internationally. Data use is governed by the Birhan Data Access Committee (DAC) and follows Birhan’s data sharing policy. All researchers who wish to access Birhan data can complete a Birhan data request form and submit it for decision by the Birhan DAC. Datasets will only be provided with de-identified data to maintain confidentiality of study participants. Requests for data can be sent to the principal investigators at. Birhan serves as the field site for HaSET (‘happiness’ in Amharic)—a maternal and child health research program in Ethiopia. For further details, please visit www.hasetmch.org.

## Profile in a nutshell

- The Birhan HDSS was established in 2018 to set up a platform for community- and facility-based research and research training. The specific objectives at inception are to generate high quality data on the health and demographic profile of a rural Ethiopia, with a specific focus on maternal and child health.
- The site is located in the North Shewa Zone of the Amhara Region, 130 km north of the capital Addis Ababa.
- The site has 18,933 households and a population of 77,766. Over 82.6% of the population is rural and includes newborns, children, adolescents, adults, and the elderly. It has a population residing in a wide range of topographic and climatic environments with diverse morbidities such as febrile illnesses, diarrheal diseases, upper respiratory infections.
- All houses are geocoded; data are collected every three months on sociodemographic status; housing construction material; economics and asset ownership; care seeking behaviors; immunization status; water and sanitation access, and key demographic events such as births, deaths, marital status changes, in-migration, or out-migration.
- Ongoing studies include pregnancy surveillance, estimating causes of mortality using verbal autopsy, determining immunization coverage, and evaluating access to water and sanitation infrastructure.

## Data Availability

All researchers who wish to access Birhan data can complete a Birhan data request form and submit it for decision by the Birhan DAC. Datasets will only be provided with de-identified data to maintain confidentiality of study participants. Requests for data can be sent to the principal investigators at. For further details, please visit www.hasetmch.org

## Acknowledgements

We are indebted to the Birhan mothers, children, and families for their ongoing participation. We acknowledge all Birhan staff and community informants who have worked tirelessly for the successful establishment of the site. We are grateful to St. Paul’s Hospital Millennium Medical College, the Amhara Regional Health Bureau, North Shewa Zone Office, and Angolela Tera and Kewet/Shewa Robit Woreda Offices for their support. Specifically, we thank Dr. Wendmagegn Gezahegn, Dr. Sisay Sirgu, and Dr. Balkachew Nigatu for their continuous support. We appreciate the advice from Dr. Robert Breiman and Dr. Nega Assefa as we established the site.

## References

1. Maternal mortality facts sheets: World Health Organization; 2019 [Available from: https://www.who.int/news-room/fact-sheets/detail/maternal-mortality.

2. EDHS. Ethiopia Demographic and Health Survey Prelminary Report 2016. Addis Ababa, Ethiopia and Calverton, Maryland: Central Statistical Authority and ORC Macro. 2016.

3. World Health Organization. Trends in maternal mortality 2000 to 2017: estimates by WHO, UNICEF, UNFPA, World Bank Group and the United Nations Population Division. 2019.

4. Levels & Trends in Child Mortality Report 2019, Estimates developed by the UN Inter-agency Group for Child Mortality Estimation. United Nations Children’s Fund; 2019.

5. Opportunities for Africa’s Newborns, Practical data, policy and programmatic support for newborn care in Africa. Dinah Lord RW, Leslie Elder, Kristina Grear, Alicia Antayhua, editor. South Africa: The Partnership For Maternal, Newborn & Child Health.

6. Ethiopia: Administrative map 2017 [Available from: https://reliefweb.int/sites/reliefweb.int/files/resources/21_adm_eth_081517_a0.pdf.

7. ArcGISOnline, cartographer Ethiopia_AdminBoundariesService Definition, Data Last Updated: Sep 4, 2015, 12:33:35 PM.

8. Ethiopian Meteorological Agency. Monthly Climatic Record of Temperature and Rainfall for Debrebirhan and Shewarobit cities. 2017.

9. INDEPTH Network. Population and Health in Developing Countries Volume 1. Population, Health, and Survival at INDEPTH Sites., IRDC, Ottawa, Canada. 2002.

10. Assefa N, Berhane Y, Worku A. Pregnancy rates and pregnancy loss in Eastern Ethiopia. Acta Obstet Gynecol Scand. 2013;92(6):642–7.

11. Boerma JT, Black RE, Sommerfelt AE, Rutstein SO, Bicego GT. Accuracy and completeness of mothers’ recall of diarrhoea occurrence in pre-school children in demographic and health surveys. Int J Epidemiol. 1991;20(4):1073–80.

12. Byass P, Hanlon PW. Daily morbidity records: recall and reliability. Int J Epidemiol. 1994;23(4):757–63.

13. Feikin DR, Audi A, Olack B, Bigogo GM, Polyak C, Burke H, et al. Evaluation of the optimal recall period for disease symptoms in home-based morbidity surveillance in rural and urban Kenya. Int J Epidemiol. 2010;39(2):450–8.

14. Ariff S, Lee AC, Lawn J, Bhutta ZA. Global Burden, Epidemiologic Trends, and Prevention of Intrapartum-Related Deaths in Low-Resource Settings. Clin Perinatol. 2016;43(3):593–608.

15. Harrison LH, Moursi S, Guinena AH, Gadomski AM, el-Ansary KS, Khallaf N, et al. Maternal reporting of acute respiratory infection in Egypt. Int J Epidemiol. 1995;24(5):1058–63.

16. Manual for the training of interviewers on the use of the 2016 WHO VA instrument Geneva: World Health Organization; 2017.

17. Open Data kit (ODK): University of Washington’s Computer Science and Engineering department; 2020 [cited 2020 August]. Available from: https://www.opendatakit.org/.

18. Population Projections for Ethiopia 2007-2037. Central Statistical Agency; July 2013.

19. EDHS. Ethiopia Mini Demographic and Health Survey 2019: Key Indicators. Rockville, Maryland, USA: EPHI and ICF: Ethiopian Public Health Institute (EPHI) [Ethiopia] and ICF; 2019.

20. Progress on drinking water, sanitation and hygiene: 2017 update and SDG baselines. Geneva: World Health Organization (WHO) and the United Nations Children’s Fund (UNICEF); 2017.

